# The Effect of Cognitive Tasks on Posture Control in Young Adults

**DOI:** 10.1101/2025.08.03.25332897

**Authors:** Lei Zhang, Qingjie Wang, Yuanyuan Ren, Aming Lu

## Abstract

**Objective:** Cognitive tasks play a pivotal role in the posture control of young adults. The study aims to explore the characteristics of static posture control under dual-task conditions by using wavelet analysis, a frequency-domain analysis method, so as to understand the dynamic changes of posture control of young adults under dual-task conditions more comprehensively.

**Methods:** 24 young adults (mean age 22.6 ± 2.3 years) completed single and cognitive-postural tasks on a force plate. Single-leg stance with eyes open lasted 60 seconds; cognitive task involved subtracting 7 from random numbers. Time-frequency features of center of pressure (COP) signal and wavelet decomposition energy were computed. Paired t-tests analyzed data.

**Results:** Dual-task group, compared to single-task, showed significant differences in COP indicators: total trajectory length, normalized length, and average velocity (*p* <0.05), notably in y-axis length and velocity (*p* <0.001). Energy content analysis found dual-task group significantly differed in energy ratio of four frequency bands along x-axis (*p* < 0.05), but no differences along y-axis.

**Conclusion:** Postural control ability in young adults was significantly impaired under dual-task conditions, manifested through reduced balance stability and altered sensory dependence strategies - specifically, increased reliance on proprioception over visual input. It provides a crucial basis for public health intervention to prevent falls, suggesting that proprioceptive training should be strengthened for high-risk groups, such as the elderly and occupationally exposed groups, and dual-task training should be incorporated into public health promotion strategies to reduce the risks related to falls.

## Introduction

Postural control is a crucial regulatory mechanism for maintaining an upright state and balance in the human body. It serves as the foundation for various age groups’ motor abilities and holds significance across different stages of life. Research has shown that static postural control ability might be relevant to certain sports performances (Jadczak et al., 2019; Matsuda et al., 2008). Thus, postural control has emerged as a significant topic in the fields of public health.

In daily life, individuals frequently engage in dual-tasking, such as pedestrians being vigilant for vehicles while crossing the road. Scholars refer to tasks that occupy cognitive resources as cognitive tasks, and the concurrent execution of two tasks by the human body is termed dual-tasking. Current research suggests that during cognitive tasks, a complex interaction between visual, vestibular, proprioceptive feedback systems, and the motor system is orchestrated to achieve postural control (Muelas Pérez et al., 2014). In recent years, the interaction between cognitive ability and postural control in the context of dual-tasking has garnered considerable attention among researchers. Tramontano et al.(Tramontano et al., 2017) have investigated the running speed of individuals performing single tasks in a dual-task mode, revealing reduced speeds for both cognitive and balance tasks, indicating mutual interference. Another study reached a similar conclusion that upright control capabilities decrease under dual-task conditions, suggesting a competition between visual control and cognitive task processing (Meng et al., 2019). It has also been observed that the difficulty level of cognitive tasks and the associated attentional demands differentially affect postural control (Talarico et al., 2017). However, several recent researchers have proposed that postural control may operate as an automated process, where conscious focus on movements can interfere with this process (Potvin-Desrochers et al., 2017; Richer & Lajoie, 2020; St-Amant et al., 2020; Walsh, 2021). The introduction of cognitive tasks might redirect attention, thereby reducing postural sway. Several studies have detected increased activation in prefrontal cortex and sensorimotor cortex through neuroimaging techniques, possibly reflecting the brain’s need to integrate more sensory information in dual-task conditions, but the results have also varied (Pan & Zhang, 2024; Rosso et al., 2017; Saraiva et al., 2023). Consequently, conclusions on the effects of dual-tasking on postural control are currently inconsistent. This contradictory result may be related to the common use of center of pressure (COP) time-domain metrics in postural control studies to assess postural control. Studies have shown that changes in COP time-domain indicators do not necessarily imply weakened or enhanced postural control (Hadadi & Abbasi, 2019; Stins et al., 2011). On the contrary, postural stabilization may indicate efficient regulatory mechanisms within the body. Fortunately, frequency-domain analysis techniques like wavelet analysis have gained popularity in postural control research in recent years. Applying wavelet analysis may help us better understand the characteristics of postural control under dual-task conditions. Wavelet analysis can decompose COP signals into different frequency components, with each component corresponding to a specific postural control system (Chagdes et al., 2009). Muscles and proprioceptive senses, the cerebellum, the vestibular system, and vision correspond to different frequency components of COP information, from high to low frequency bands (Chagdes et al., 2009; Oppenheim et al., 1999; Paillard et al., 2002; Quek et al., 2014). Wavelet analysis seems to provide a more nuanced assessment of balance function in specific populations and postural recovery scenarios (Lacour et al., 2021; Liang et al., 2014; Quek et al., 2014).

Thus, this study aims to provide a detailed characterization of static postural control under dual-task conditions using wavelet analysis. We hypothesized that dual-tasking would increase postural instability and that cognitive tasks would alter frequency band energy, thereby inducing changes in sensory integration and motor control strategies. A thorough understanding of the mechanisms through which cognitive load impairs postural control (e.g., increased reliance on proprioception versus the visual system) could offer a scientific basis for developing public health interventions to prevent falls.

## Research Methods

### Participants

This study was approved by the Research Ethics Committee of Soochow University (No. ECSU-2019000209). Sample size was calculated using G*power software. Referring to previous study (Y. Ren et al., 2022), setting an effect size of 0.8, α = 0.05, and power = 0.95 yielded a minimum sample size of 19. Considering the possible sample loss rate, 24 participants were included (mean age: 22.6 ± 2.3 years, mean height: 174.9 ± 8.4 cm, mean weight: 70.1 ± 10.9 kg). All participants exhibited normal cognitive function, with no history of neurological or muscular impairments. Additionally, none had sustained lower limb joint injuries within the six months preceding the study, and all refrained from vigorous physical activity for at least 24 hours prior to the experiment.

### Experimental Setup

The experiment employed a Swiss-made KISTLER three-dimensional force plate, model 9281A, with dimensions of 60 cm × 40 cm (Figure 1). The center of pressure (COP) signal was derived from the ground reaction force (GRF) data collected by the force plate at a sampling frequency of 1000 Hz. The data was exported through a Datalog data box.

**Fig 1.**
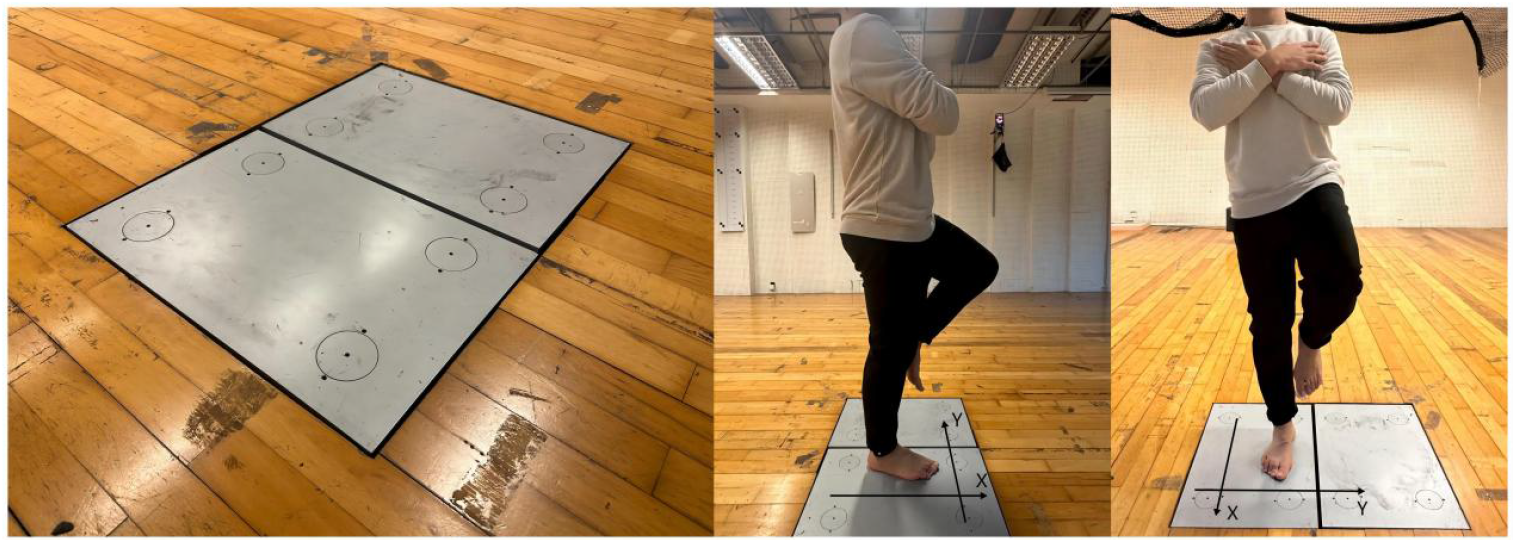
KISTLER three-dimensional force plate and one leg stand test

### Experimental Procedure

The testing process consisted of two parts: single-task postural control (ST) and cognitive-postural dual-task (DT). Each participant completed both tasks twice, with the order randomized and a resting period of at least 1 minute between tasks. Before the test commenced, participants were informed that they should prioritize completing the cognitive task. For the single-task condition, participants were instructed to stand on one leg (dominant side), barefoot, on the force plate. They fixated their gaze on a black dot on a whiteboard located 2 meters away, crossing their arms over their chests and maintaining a single-leg stance for 60 seconds (Figure 1). Participants aimed to maintain an upright position with minimal sway. In the dual-task condition, participants simultaneously performed the postural control task and the cognitive task. The postural control task was the same as the single-task condition. According to previous literature, the cognitive task chosen was an arithmetic task (Chen & Bailey, 2021; Lemaire, 2021; Xu et al., 2023). Participants mentally subtracted 7 from a randomly chosen number between 300 and 900, and errors were not corrected during the task.

### Data Processing

The raw COP signal was resampled to 100 Hz. Research has shown that 95% of the COP signal is concentrated below 6.25 Hz (Thomas et al., 2018). The original COP signal was filtered using a 2nd-order Butterworth low pass filter with a cutoff frequency of 12.5 Hz. MATLAB was used for data analysis. Time-domain indicators included total trajectory length Lxy, x-axis trajectory length Lx, y-axis trajectory length Ly, unit area trajectory length Lxy/Area, envelope area Area, average velocity Vxy, x-axis average velocity Vx, y-axis average velocity Vy. The frequency domain processing adopts 9-level Symlet-8 discrete wavelet transform, and the signal reconstruction process is shown in Figure 2, which decomposes the original signal layer by layer. Each decomposition divides the the signal is divided into low-frequency and high-frequency parts. Immediately after this, the low frequency signal obtained from the decomposition is again decomposed to obtain two parts, for a total of nine times. A1, A2……A9 are low-frequency signals and D1, D2……D8 are high-frequency signals. As Figure 3 showed, the COP data was decomposed into four frequency bands: moderate frequency (1.56-6.25 Hz), low frequency (0.39-1.56 Hz), very low frequency (0.10-0.39 Hz) and ultra-low frequency (<0.10 Hz) (Wang et al., 2021). The energy content of each frequency band to the total COP signal was calculated.

**Fig 2.**
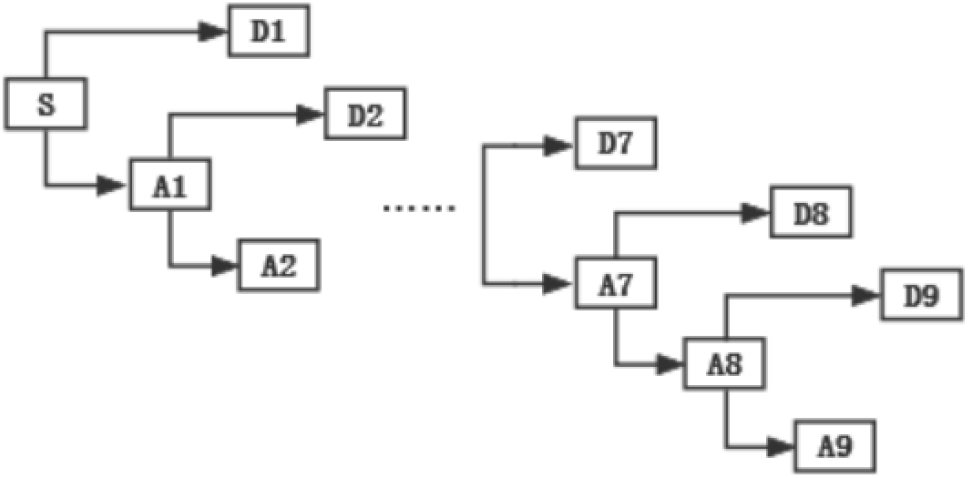
Flowchart of wavelet cascade decomposition. Notes: The original COP signal is decomposed layer by layer into high frequency and low frequency parts by 9-level Symlet-8 discrete wavelet. S is the original COP signal, A is the low frequency signal and D is the high frequency signal.

**Fig 3.**
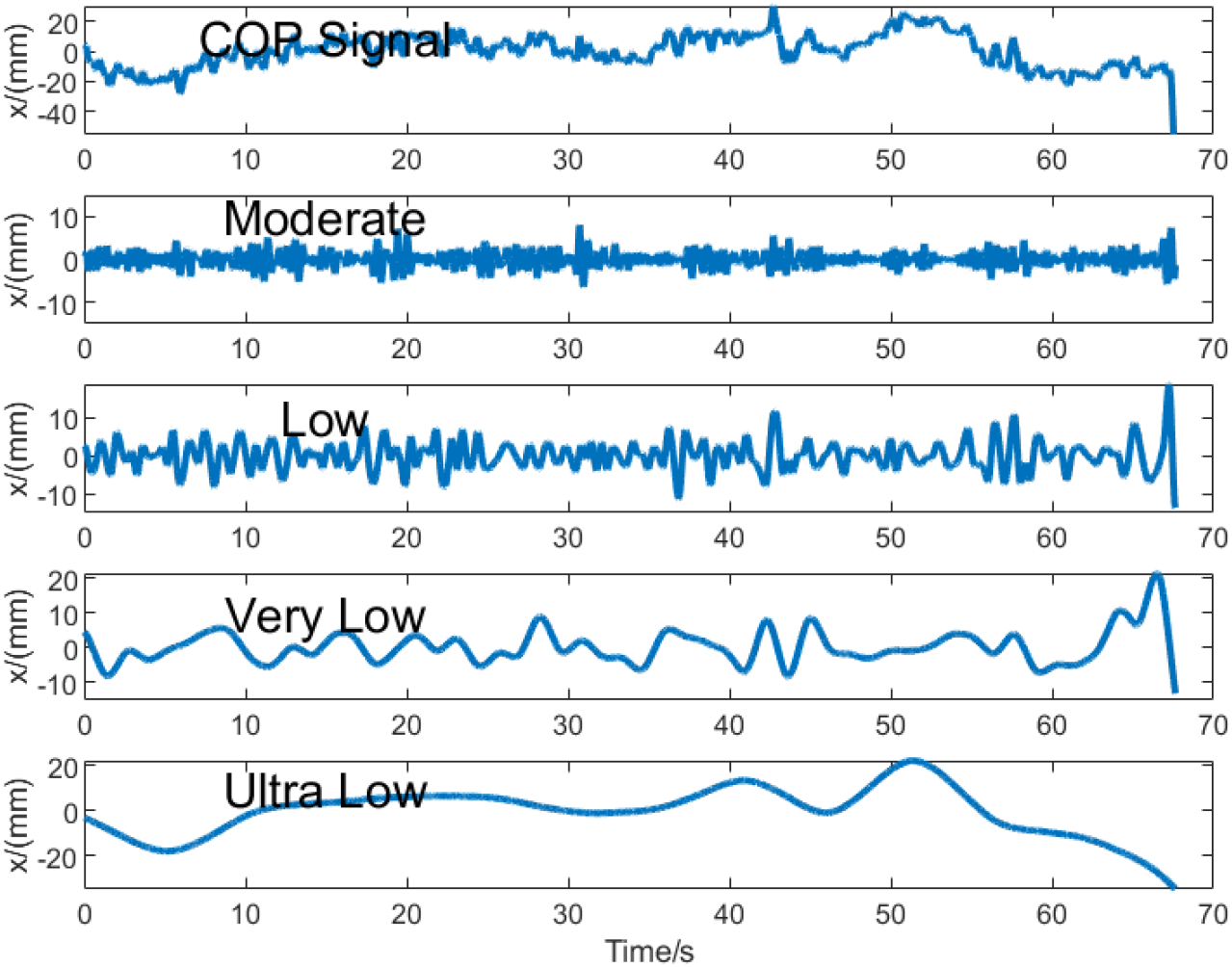
COP original signal and reconstructed band signal. Notes: moderate frequency (1.56-6.25 Hz), low frequency (0.39-1.56 Hz), very low frequency (0.10-0.39 Hz) and ultra low frequency (<0.10 Hz).

The timing energy of each frequency band is calculated with the following formula:

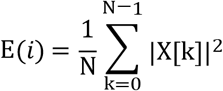

E_(*i*)_is the total energy of frequency band i of the COP signal. N is the total amount of data in frequency band a. X is the temporal data set, and k denotes the kth data in X set.

The ratio of energy to total energy for each frequency band is calculated with the following formula:

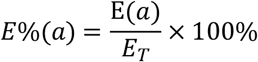

*E*%(*a*) denotes the percentage of the COP signal a-band energy in the total energy. E(*a*) is the total COP signal a-band energy, *E*_*T*_ and is the total COP signal energy.

### Statistical Analysis

All data were analyzed using SPSS 26.0 and presented as means ± standard deviations (M ± SD). Initially, normality tests were conducted on both single-task and dual-task data. When the data satisfied the normal distribution assumption, paired sample t-tests were used for significance analysis. The statistical significance level was set at *p* < 0.05, with *p* < 0.001 indicating a highly significant difference.

## Results

### COP Indicators

As with table 1 and figure 4 displays, compared with the single-task condition, the dual-task condition showed significant differences in Lxy, Lxy/Area, and Vxy (*p* <0.05), and highly significant differences in Ly and Vy (*p* <0.001). The dual-task condition demonstrated a smaller Lxy and Lxy/Area, indicating a longer COP trajectory length and bigger area covered during the single-leg stance, respectively. Additionally, the Vxy of the dual-task condition was higher than that of the single-task condition, suggesting increased velocity.

**Table 1.** envelope area related indicator test results.

| Indicators | Group | M±SD | *t* | *p* |
| --- | --- | --- | --- | --- |
| Area(mm<sup>2</sup>) | ST | 472.21±145.22 | 1.592 | 0.128 |
| | DT | 398.24±130.20 | | |
| $L_{xy}/Area(mm^{-1})$ | ST | 9.65±3.32 | -2.662 | 0.015* |
| | DT | 12.31±3.61 | | |

Note: “\*” means $p < 0.05$ , “\*\*\*” means $p < 0.001$ ; ST, single task; DT, dual task.

**Fig 4.**
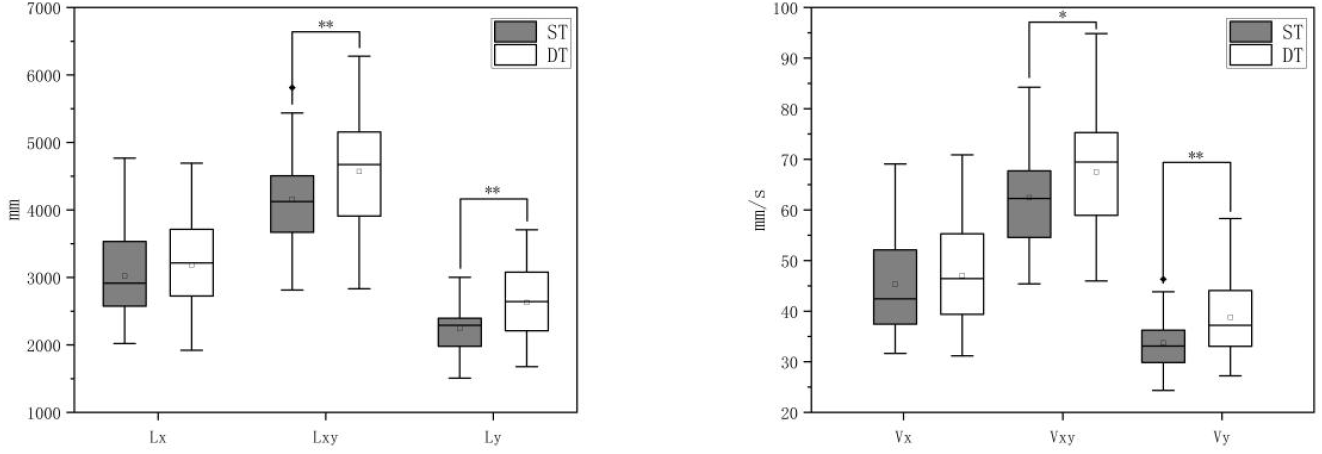
box diagram of trajectory lengths and velocity. Note: “*”means *p* <0.05, “**”means *p* <0.001; ST, single task; DT, dual task.

### Energy content

The energy content of different frequency bands in both the x and y directions is presented in Figure 5. In the x-axis direction, significant differences were observed in the four frequency bands between the single-task and dual-task conditions (*p* <0.05), with the dual-task condition demonstrating lower energy content in the moderate, low, and very low frequency bands but higher energy content in the ultra low frequency band. However, in the y-axis direction, no significant differences were observed in the energy content of the four frequency bands between the single-task and dual-task conditions.

**Fig 5.**
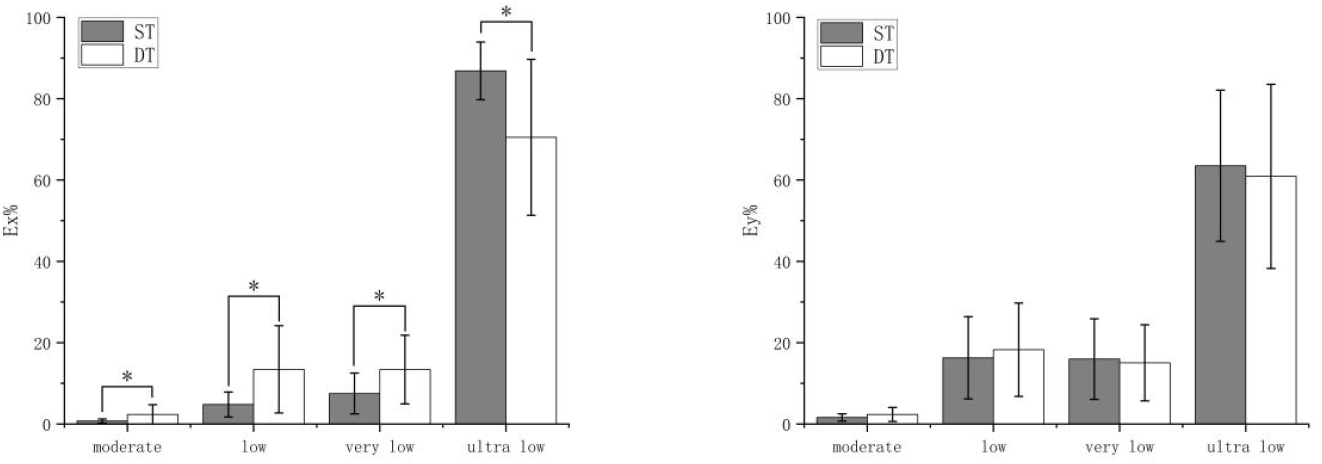
COP band energy comparison. Note: “*”means *p* <0.05; ST, single task; DT, dual task.

## Discussion

The results indicated significant differences between the single-task and dual-task groups in the total trajectory length Lxy, unit area trajectory length Lxy/Area, average velocity Vxy, y-axis trajectory length Ly, and average velocity Vy. Changes in the total trajectory length suggest a decrease in postural control stability under dual-task conditions. The differences in unit area trajectory length may indicate changes in the complexity or interference level of postural control. Alterations in the overall average velocity reflect dynamic changes in postural control, suggesting increased complexity under dual-task conditions. Additionally, the y-axis trajectory length and average velocity provide detailed information on postural control in the y-axis direction, highlighting changes in postural fluctuations and adjustment speed along this axis. Overall, these changes suggest that dual-task conditions have a comprehensive impact on postural control, potentially indicating a significant reduction in postural control capability. Postural control involves sensory input integration and motor output; its impairment likely stems from these three facets (Beurskens et al., 2016). Sensory input relies on vestibular, proprioceptive, and visual cues (Alizadeh et al., 2022).

While this study didn’t directly validate the impact of these sensory inputs on postural control, frequency-domain information can infer control mechanisms and system involvement levels. Typically, a faster neuromuscular response corresponds to a higher frequency. Medium frequencies, on the other hand, reflect muscular and proprioceptive actions. In contrast, the vestibular system and visual modulation are slow and stable, with lower frequencies. Thus, wavelet-analyzed COP signals represent various factors: muscle proprioception, neural system, vestibular, and visual, aligned with different frequency bands (Chagdes et al., 2009; Oppenheim et al., 1999; Paillard et al., 2002; Quek et al., 2014). Frequency band energy content reveals in the dual-task group, compared to the single-task group, ultra-low-frequency energy decreased in the anterior-posterior direction (x-axis), while other frequency bands increased. However, energy content in the left-right direction (y-axis) showed no significant difference. This indicates that the cognitive task may have led to a reallocation of CNS resources and a greater degree of interference with postural control. Ultra-low-frequency energy reduction suggests a weakened visual role in postural control (Quek et al., 2014; Wang et al., 2021); increased energy in other frequency bands indicates heightened reliance on proprioceptive and vestibular systems (Chagdes et al., 2009). Reduced visual input causes increased body sway, accompanied by active and passive muscle reflexes (Cheung & Schmuckler, 2021). When the head moves, vestibular system amplification maintains head stability, and muscle-joint activities transmit proprioceptive signals to the nervous system (Chepisheva, 2023; Cone et al., 2017). Despite augmented vestibular and proprioceptive influences, they cannot fully compensate for visual deficits, leading to an overall decrease in stability. In studies linking cognition and postural control, results have shown inconsistent congruity. Some researchers find cognitive tasks have no impact (Yardley et al., 1999), or even a facilitatory effect (Stins et al., 2011), likely due to task complexity (Barra et al., 2006). Increased cognitive task difficulty influences balance performance, affecting tasks of varying difficulty levels (Ruffieux et al., 2015). Task complexity changes attention allocation (Boisgontier et al., 2013), with increased difficulty potentially consuming more attention resources and impacting task performance (Woollacott & Shumway-Cook, 2002). Additionally, divergent brain region activation due to distinct cognitive task types and cognitive task enhancement might play roles.

The integrated function of the nervous system is pivotal for maintaining balance. In this study, an increase in low-frequency energy content in the anterior-posterior direction indicates heightened nervous system activity. Research on the neural mechanisms during dual-task conditions is abundant. Currently, two theories are widely accepted for the decline in performance during dual tasks: the capacity-sharing model (Tombu & Jolicoeur, 2003) and the bottleneck (task-switching) model (Pashler, 1994). We can also understand them as parallel and serial paradigms. A single theoretical model seems difficult to explain all task paradigms (I et al., 2018). It has been shown that when performing dual tasks, the human body is free to switch between serial and parallel, but prefers parallel processing, also known as capacity sharing mode (C & R, 2009). Some researchers, employing functional near-infrared spectroscopy (fNIRS), observed significant activation in the prefrontal cortex during dual tasks (Basso Moro et al., 2014). This suggests a significant role of the prefrontal cortex in both cognitive and postural control dual tasks, regardless of the theoretical framework. The prefrontal cortex is regarded as responsible for attention allocation (Holtzer et al., 2015; Mihara et al., 2008) and processing visual and proprioceptive information (Ferrari et al., 2020). Additionally, the dorsolateral prefrontal cortex is suggested to manage memory tasks (Barbey et al., 2013; McCarthy et al., 1994; Petrides, 2000). Researchers like Ren Jie (J. Ren et al., 2010) investigated cognitive task effects on visual control during standing posture, finding that cognitive tasks inhibit visual function and highlight the shared central processing system for cognition and balance. This implies a cognitive-motor conflict in the brain during the execution of cognitive and balance tasks, indirectly explaining reduced visual effects and diminished postural control capability as observed in this study. Moreover, the increase in moderate-frequency energy suggests heightened involvement of muscles and proprioception in postural control. During cognitive tasks, muscle tension increases in an upright stance to minimize limb swing and joint movement. Vestibular and proprioceptive signals transmit balance-related information to the central system, facilitating timely adjustments in the motor system. However, due to weakened visual effects, the adjustment process is slower, leading to increased muscle activity.

The innovation of this study lies in its application of wavelet analysis to investigate postural control under dual-task conditions. By conducting a detailed time-frequency domain analysis, this study provides deeper insights into the postural control mechanisms of young adults across different frequency bands during cognitive tasks. These findings offer a scientific basis for developing targeted public health interventions to prevent falls. However, there are still some limitations. First, the study only investigated young adults, but age and gender are also important influences, so future research can continue to discuss the effects of dual-tasking on other groups such as women, children and the elderly. Second, the cognitive task used in this study involved mental subtraction, although it serves the purpose of dual-tasking, it does not fully reflect the diversity of cognitive tasks that individuals face in the real world. Future studies could investigate a wider range of cognitive tasks.

## Conclusion

Under dual-task conditions, young adults exhibit significantly compromised postural control, characterized by diminished balance stability and a shift in sensory integration strategies from visual to proprioceptive dominance. These findings have substantial public health relevance, underscoring the negative impact of contemporary multitasking demands on postural stability. Importantly, the study demonstrates that cognitive load shifts sensory dependence from visual to proprioceptive systems. This critical insight provides scientific justification for implementing targeted proprioception training in fall prevention programs, particularly for occupational groups(e.g., healthcare professionals and firefighters) who routinely engage in cognitively demanding tasks while maintaining physical stability.

## Data Availability

All relevant data are within the manuscript and its Supporting Information files.

## Reference

Alizadeh, A., Jafarpisheh, A. S., Mohammadi, M., & Kahlaee, A. H. (2022). Visual, Vestibular, and Proprioceptive Dependency of the Control of Posture in Chronic Neck Pain Patients. Motor Control, 26(3), 362–377. 10.1123/mc.2021-0008

Barbey, A. K., Koenigs, M., & Grafman, J. (2013). Dorsolateral prefrontal contributions to human working memory. Cortex; a Journal Devoted to the Study of the Nervous System and Behavior, 49(5), 1195–1205. 10.1016/j.cortex.2012.05.022

Barra, J., Bray, A., Sahni, V., Golding, J. F., & Gresty, M. A. (2006). Increasing cognitive load with increasing balance challenge: Recipe for catastrophe. Experimental Brain Research, 174(4), 734–745. 10.1007/s00221-006-0519-2

Basso Moro, S., Bisconti, S., Muthalib, M., Spezialetti, M., Cutini, S., Ferrari, M., Placidi, G., & Quaresima, V. (2014). A semi-immersive virtual reality incremental swing balance task activates prefrontal cortex: A functional near-infrared spectroscopy study. NeuroImage, 85 Pt 1, 451–460. 10.1016/j.neuroimage.2013.05.031

Beurskens, R., Haeger, M., Kliegl, R., Roecker, K., & Granacher, U. (2016). Postural Control in Dual-Task Situations: Does Whole-Body Fatigue Matter? PloS One, 11(1), e0147392. 10.1371/journal.pone.0147392

Boisgontier, M. P., Beets, I. A. M., Duysens, J., Nieuwboer, A., Krampe, R. T., & Swinnen, S. P. (2013). Age-related differences in attentional cost associated with postural dual tasks: Increased recruitment of generic cognitive resources in older adults. Neuroscience and Biobehavioral Reviews, 37(8), 1824–1837. 10.1016/j.neubiorev.2013.07.014

C, L., & R, H. (2009). Strategic capacity sharing between two tasks: Evidence from tasks with the same and with different task sets. Psychological Research, 73(5). 10.1007/s00426-008-0162-6

Chagdes, J. R., Rietdyk, S., Haddad, J. M., Zelaznik, H. N., Raman, A., Rhea, C. K., & Silver, T. A. (2009). Multiple timescales in postural dynamics associated with vision and a secondary task are revealed by wavelet analysis. Experimental Brain Research, 197(3), 297–310. 10.1007/s00221-009-1915-1

Chen, E. H., & Bailey, D. H. (2021). Dual-task studies of working memory and arithmetic performance: A meta-analysis. Journal of Experimental Psychology. Learning, Memory, and Cognition, 47(2), 220–233. 10.1037/xlm0000822

Chepisheva, M. K. (2023). Spatial orientation, postural control and the vestibular system in healthy elderly and Alzheimer’s dementia. PeerJ, 11, e15040. 10.7717/peerj.15040

Cheung, T. C. K., & Schmuckler, M. A. (2021). Multisensory postural control in adults: Variation in visual, haptic, and proprioceptive inputs. Human Movement Science, 79, 102845. 10.1016/j.humov.2021.102845

Cone, B. L., Goble, D. J., & Rhea, C. K. (2017). Relationship between changes in vestibular sensory reweighting and postural control complexity. Experimental Brain Research, 235(2), 547–554. 10.1007/s00221-016-4814-2

Ferrari, N., Schmitz, L., Schmidt, N., Mahabir, E., Van de Vondel, P., Merz, W. M., Lehmacher, W., Stock, S., Brockmeier, K., Ensenauer, R., Fehm, T., & Joisten, C. (2020). A lifestyle intervention during pregnancy to reduce obesity in early childhood: The study protocol of ADEBAR - a randomized controlled trial. BMC Sports Science, Medicine & Rehabilitation, 12, 55. 10.1186/s13102-020-00198-5

Hadadi, M., & Abbasi, F. (2019). Comparison of the Effect of the Combined Mechanism Ankle Support on Static and Dynamic Postural Control of Chronic Ankle Instability Patients. Foot & Ankle International, 40(6), 702–709. 10.1177/1071100719833993

Holtzer, R., Mahoney, J. R., Izzetoglu, M., Wang, C., England, S., & Verghese, J. (2015). Online fronto-cortical control of simple and attention-demanding locomotion in humans. NeuroImage, 112, 152–159. 10.1016/j.neuroimage.2015.03.002

I, K., E, P., H, M., & A, K. (2018). Cognitive structure, flexibility, and plasticity in human multitasking-An integrative review of dual-task and task-switching research. Psychological Bulletin, 144(6). 10.1037/bul0000144

Jadczak, Ł., Grygorowicz, M., Wieczorek, A., & Śliwowski, R. (2019). Analysis of static balance performance and dynamic postural priority according to playing position in elite soccer players. Gait & Posture, 74, 148–153. 10.1016/j.gaitpost.2019.09.008

Lacour, M., Tardivet, L., & Thiry, A. (2021). Posture Deficits and Recovery After Unilateral Vestibular Loss: Early Rehabilitation and Degree of Hypofunction Matter. Frontiers in Human Neuroscience, 15, 776970. 10.3389/fnhum.2021.776970

Lemaire, P. (2021). Effects of prior-task failure on arithmetic performance: A study in young and older adults. Memory & Cognition, 49(6), 1236–1246. 10.3758/s13421-021-01161-6

Liang, Z., Clark, R., Bryant, A., Quek, J., & Pua, Y. H. (2014). Neck musculature fatigue affects specific frequency bands of postural dynamics during quiet standing. Gait & Posture, 39(1), 397–403. 10.1016/j.gaitpost.2013.08.007

Matsuda, S., Demura, S., & Uchiyama, M. (2008). Centre of pressure sway characteristics during static one-legged stance of athletes from different sports. Journal of Sports Sciences, 26(7), 775–779. 10.1080/02640410701824099

McCarthy, G., Blamire, A. M., Puce, A., Nobre, A. C., Bloch, G., Hyder, F., Goldman-Rakic, P., & Shulman, R. G. (1994). Functional magnetic resonance imaging of human prefrontal cortex activation during a spatial working memory task. Proceedings of the National Academy of Sciences of the United States of America, 91(18), 8690–8694. 10.1073/pnas.91.18.8690

Meng, H.-J., Luo, S.-S., & Wang, Y.-G. (2019). The interplay between cognitive tasks and vision for upright posture balance in adolescents. PeerJ, 7, e7693. 10.7717/peerj.7693

Mihara, M., Miyai, I., Hatakenaka, M., Kubota, K., & Sakoda, S. (2008). Role of the prefrontal cortex in human balance control. NeuroImage, 43(2), 329–336. 10.1016/j.neuroimage.2008.07.029

Muelas Pérez, R., Sabido Solana, R., Barbado Murillo, D., & Moreno Hernández, F. J. (2014). Visual availability, balance performance and movement complexity in dancers. Gait & Posture, 40(4), 556–560. 10.1016/j.gaitpost.2014.06.021

Oppenheim, U., Kohen-Raz, R., Alex, D., Kohen-Raz, A., & Azarya, M. (1999). Postural characteristics of diabetic neuropathy. Diabetes Care, 22(2), 328–332. 10.2337/diacare.22.2.328

Paillard, T., Costes-Salon, C., Lafont, C., & Dupui, P. (2002). Are there differences in postural regulation according to the level of competition in judoists? British Journal of Sports Medicine, 36(4), 304–305. 10.1136/bjsm.36.4.304

Pan, J., & Zhang, S. (2024). Dual-Task Effect on Center of Pressure Oscillations and Prefrontal Cortex Activation Between Young and Older Adults. Research Quarterly for Exercise and Sport, 1–11. 10.1080/02701367.2024.2365940

Pashler, H. (1994). Dual-task interference in simple tasks: Data and theory. Psychological Bulletin, 116(2), 220–244. 10.1037/0033-2909.116.2.220

Petrides, M. (2000). The role of the mid-dorsolateral prefrontal cortex in working memory. Experimental Brain Research, 133(1), 44–54. 10.1007/s002210000399

Potvin-Desrochers, A., Richer, N., & Lajoie, Y. (2017). Cognitive tasks promote automatization of postural control in young and older adults. Gait & Posture, 57, 40–45. 10.1016/j.gaitpost.2017.05.019

Quek, J., Brauer, S. G., Clark, R., & Treleaven, J. (2014). New insights into neck-pain-related postural control using measures of signal frequency and complexity in older adults. Gait & Posture, 39(4), 1069–1073. 10.1016/j.gaitpost.2014.01.009

Ren, J., Kazuhiko, W., & Makoto, M. (2010). The Effect of Cognitive Task on Visual Control of Standing Posture. Acta Psychologica Sinica, 42(3), 360–366.

Ren, Y., Wang, C., Zhang, L., & Lu, A. (2022). The effects of visual cognitive tasks on landing stability and lower extremity injury risk in high-level soccer players. Gait & Posture, 92, 230–235. 10.1016/j.gaitpost.2021.11.031

Richer, N., & Lajoie, Y. (2020). Automaticity of Postural Control while Dual-tasking Revealed in Young and Older Adults. Experimental Aging Research, 46(1), 1–21. 10.1080/0361073X.2019.1693044

Rosso, A. L., Cenciarini, M., Sparto, P. J., Loughlin, P. J., Furman, J. M., & Huppert, T. J. (2017). Neuroimaging of an attention demanding dual-task during dynamic postural control. Gait & Posture, 57, 193–198. 10.1016/j.gaitpost.2017.06.013

Ruffieux, J., Keller, M., Lauber, B., & Taube, W. (2015). Changes in Standing and Walking Performance Under Dual-Task Conditions Across the Lifespan. Sports Medicine (Auckland, N.Z.), 45(12), 1739–1758. 10.1007/s40279-015-0369-9

Saraiva, M., Castro, M. A., & Vilas-Boas, J. P. (2023). Muscular and Prefrontal Cortex Activity during Dual-Task Performing in Young Adults. European Journal of Investigation in Health, Psychology and Education, 13(4), 736–747. 10.3390/ejihpe13040055

St-Amant, G., Rahman, T., Polskaia, N., Fraser, S., & Lajoie, Y. (2020). Unveilling the cerebral and sensory contributions to automatic postural control during dual-task standing. Human Movement Science, 70, 102587. 10.1016/j.humov.2020.102587

Stins, J. F., Roerdink, M., & Beek, P. J. (2011). To freeze or not to freeze? Affective and cognitive perturbations have markedly different effects on postural control. Human Movement Science, 30(2), 190–202. 10.1016/j.humov.2010.05.013

Talarico, M. K., Lynall, R. C., Mauntel, T. C., Weinhold, P. S., Padua, D. A., & Mihalik, J. P. (2017). Static and dynamic single leg postural control performance during dual-task paradigms. Journal of Sports Sciences, 35(11), 1118–1124. 10.1080/02640414.2016.1211307

Thomas, K. S., Hammond, M., & Magal, M. (2018). Graded forward and backward walking at a matched intensity on cardiorespiratory responses and postural control. Gait & Posture, 65, 20–25. 10.1016/j.gaitpost.2018.06.168

Tombu, M., & Jolicoeur, P. (2003). A central capacity sharing model of dual-task performance. Journal of Experimental Psychology. Human Perception and Performance, 29(1), 3–18. 10.1037//0096-1523.29.1.3

Tramontano, M., Morone, G., Curcio, A., Temperoni, G., Medici, A., Morelli, D., Caltagirone, C., Paolucci, S., & Iosa, M. (2017). Maintaining gait stability during dual walking task: Effects of age and neurological disorders. European Journal of Physical and Rehabilitation Medicine, 53(1), 7–13. 10.23736/S1973-9087.16.04203-9

Walsh, G. S. (2021). Visuomotor control dynamics of quiet standing under single and dual task conditions in younger and older adults. Neuroscience Letters, 761, 136122. 10.1016/j.neulet.2021.136122

Wang, G., Mao, X., Zhang, Q., & Lu, A. (2021). Time-frequency Analysis of COP in Static Postural Control of Subjects with Functional Ankle Instability. Chinese Journal of Sports Medicine, 40(09), 691–697. 10.16038/j.1000-6710.2021.09.003

Woollacott, M., & Shumway-Cook, A. (2002). Attention and the control of posture and gait: A review of an emerging area of research. Gait & Posture, 16(1), 1–14. 10.1016/S0966-6362(01)00156-4

Xu, Y., Geng, C., Tang, T., Huang, J., & Hou, Y. (2023). How to prevent cognitive overload in the walking-arithmetic dual task among patients with Parkinson’s disease. BMC Neurology, 23(1), 205. 10.1186/s12883-023-03231-5

Yardley, L., Gardner, M., Leadbetter, A., & Lavie, N. (1999). Effect of articulatory and mental tasks on postural control. Neuroreport, 10(2), 215–219. 10.1097/00001756-199902050-00003

